# Combined accelerometry and writing analysis for quantifying tremor during focused ultrasound thalamotomy

**DOI:** 10.1101/2025.07.29.25332395

**Authors:** Andrew E. Toader, Nemanja Useinovic, Beck Shafie, Matthew C. Henn, Molly Joyce, Haley D. Smith, Lee E. Neilson, Delaram Safarpour, Ahmed M. Raslan, Daniel C. Cleary

## Abstract

**Background:** MRI-guided focused ultrasound (MRgFUS) thalamotomy treats tremors in essential tremor (ET). Identifying the correct area to ablate relies on subjective physician assessment of tremor improvement during treatment. To address this limitation, objective quantification of tremor severity during MRgFUS using an MRI-compatible accelerometer and tablet was performed.

**Objective:** To develop and evaluate an objective method for quantifying tremor progression during MRgFUS.

**Methods:** Forty patients undergoing MRgFUS thalamotomy for control of tremor were evaluated during the procedure using analysis of drawn Archimedean spirals and a pen-mounted accelerometer. The severity of the tremor was determined by analysis of the rhythmic oscillatory patterns present in the drawings and accelerometer recordings after each sonication delivered. The patient’s drawings were evaluated by two movement disorder neurologists, using the drawing subsection of The Essential Tremor Rating Scale (TETRAS), and subsequently compared to the TETRAS score.

**Results:** The average improvements in tremor after MRgFUS thalamotomy measured by our methods were: 63.6±7% by accelerometer analysis, 72.4±9.9% by written spiral analysis, 68±6.7% by combining the accelerometer and written spiral analysis, and 33.6±3.1% by TETRAS analysis (mean ± SEM). Stratifying improvement on the basis of temperature shows improvement of 22.8±4.4% at <50°C, 48.9±5.5% at 50-53 °C, and 70.8±2.9% at >53°C.

**Conclusions:** The performance of accelerometer and written spiral analysis shows a trend comparable to human analysis of the spirals but had greater sensitivity for subtle changes. This system of analysis provides an objective and instantaneous measure of tremor improvement during MRgFUS, potentially making the procedure safer and more efficient.

## Introduction

Up to 8% of the global population over the age of 60 has benign essential tremor (ET), making it the most common movement disorder in humans (1, 2). Severe ET reduces quality of life, causes social isolation and loss of independence, and impairs activities of daily living, self-care, and feeding (3–6). Although medications and lifestyle adjustments are effective in many cases (7, 8), approximately 50% of patients experience inadequate relief or intolerable side effects, prompting surgical treatment. In 2016, the FDA approved Magnetic Resonance Imaging (MRI)-guided high-intensity focused ultrasound (MRgFUS) thalamotomy as an option for treatment of ET. Since then, this thermo-ablative procedure has become a prominent surgical treatment modality for medication-refractory ET, with tens of thousands of patients successfully treated to date (9). Treatment with MRgFUS does not require anesthesia or incisions, includes no implanted hardware, eliminates the risk of surgical site infections, and requires no long-term follow-up programming or surgery for battery changes.

The primary target for thalamotomy is the ventral intermediate nucleus (VIM) of the thalamus, a relay point for fiber pathways from the brainstem to the primary motor cortex. The VIM is a relatively small surgical target (∼4 mm diameter) and is closely bounded by the internal capsule (laterally) and principal sensory nucleus of the thalamus (posteriorly) (10). Because of its proximity to these key structures, VIM thalamotomy carries significant risk for short- and long-term off-target effects, including weakness, numbness, and ataxia (11, 12). Off-target effects are common because the VIM cannot be directly identified on MRI; instead, stereotaxic coordinates and indirect targeting are used to approximate the location of VIM. After a starting point is determined, physiological mapping with a low-energy treatment (sonication) causes a transient functional deficit in the affected regions. A physical exam is used to assess the physiological consequences of that deficit, both the degree of tremor improvement and the presence of off-target effects. The target location is then adjusted based on the degree of tremor improvement and the nature and severity of off-target effects.

Physiological mapping in MRgFUS is effective for target confirmation but the technique is limited by the progressively increasing energy requirements in subsequent sonications (13, 14). After the initial low-energy sonication, if the degree of tremor improvement is not as expected or off-target effects are seen, then the target must be adjusted and the low dose sonication repeated. However, with repeated sonications the skull displays progressively increasing resistance to ultrasound energy, and each successive sonication affects a larger area of brain parenchyma, with increased risk of temporary or permanent off-target effects (15–18). For some patients, the clinician may only have one or two low-energy mapping trials before they must decide on a final target location. Much weighs on the clinician’s perception of subtle improvements in the patient’s tremor, which can be affected by many intrinsic and extrinsic factors (19). Accurate assessment of tremor improvement is critical for timely targeting adjustment, in order to achieve optimal treatment outcomes and minimize off-target side effects (15, 16). Adding to this complexity is that tremor response to early, low-energy sonication trials is often subtle, and quantitative measure of improvement could aid in achieving precise stereotaxic positioning and alignment.

The most common method for assessing tremor improvement is the “spiralgram”, a well-validated writing task where patients are instructed to draw an Archimedean spiral, and tremors can be visualized by repeated deviations of the drawn line from the intended path (20, 21). The Essential Tremor Rating Assessment Scale (TETRAS) is a standardized assessment method which quantifies the severity of tremor via spiralgram using an ordinal 5-point scale, but TETRAS scoring lacks the granularity to reliably detect small improvements in tremor severity (22–24).

Given that the success of MRgFUS is contingent upon the physician’s judgment of the adequacy of tremor improvement, a need exists for an objective, quantitative measure of tremor improvement, to aid in achieving optimal procedure outcomes.

To address this need, an MRI compatible analysis method was developed to more precisely capture and quantify tremor characteristics during MRgFUS. This method tracks patient hand movements with a portable accelerometer and also captures handwriting with a digital spiralgram, and then uses custom software to provide an objective, real-time measurement of tremor improvement. This study demonstrates the feasibility and performance of digital tremor analysis during patient treatment with MRgFUS. We found that this method provides objective and easily interpretable data on the state of tremors, enabling physicians to make more accurate tremor assessments and judgements on targeting adjustments earlier in the procedure.

## Methods

### Patient Demographics

A total of 40 patients with either ET or tremor dominant Parkinson’s Disease (PD) who underwent MRgFUS at a single institution from April 10th, 2024 to July 31st, 2024 were included in this study. Patient’s provided written and verbal consent to participate in IRB-approved research (IRB #00026630). Patients had a diagnosis of either essential tremor (n=30) or Tremor-Dominant Parkinson’s Disease (n=10). A summary of patient demographics is shown in table 1.

**Table 1.** Patient demographics.

| Variable | Treatment group, n (%) |
| --- | --- |
| Age, mean (range) | 74.5 (57-88) |
| Sex |  |
| Male | 30 (75%) |
| Female | 10 (25%) |
| Race/Ethnicity |  |
| White/Non-Hispanic | 37 (92.5%) |
| Declined to answer | 3 (7.5%) |
| Primary Diagnosis |  |
| Essential Tremor | 30 (75%) |
| Parkinson's Disease | 10 (25%) |
| Hemisphere Treated |  |
| Left | 29 (72.5%) |
| Right | 11 (27.5%) |
| Previous Treatment |  |
| First Treatment | 34 (85%) |
| Second Treatment, other side | 4 (10%) |
| Second Treatment, redo same side | 2 (5%) |
| Skull Density Ratio, mean (SD) | 0.54 (0.09) |

### Data Collection

To quantify tremor severity, we combined custom software for handwriting analysis with an accelerometer attached to a pen. The accelerometer is a commercially available Bluetooth-enabled motion sensor (Mbientlabs, San Jose, CA, USA), which was encased in a custom 3D-printed enclosure that attaches to a pen or stylus. Data download and analysis was performed using a python application running on Microsoft Windows based Surface Go 3 tablet (Microsoft, Redmond, WA, USA). A 3D-printed holder was attached to the back of the tablet, which allowed patients on the MRI table to have easy access to the front of the tablet. Complete instructions for installation, code for the program, and 3D-models are freely available via GitHub (https://github.com/clearylabOHSU/tremor_analysis). The patients were instructed to draw spirals with the accelerometer-connected stylus, which then transmitted the movements to the software on the tablet. The case for the accelerometer also had a hook on the side, and so could be easily attached to a cup or other object, enabling use in specific task-based assessments such as drinking from a cup. The tablet’s software displays the movement analysis within seconds, allowing the surgeon to visualize the improvement instantaneously. Our choice of tablet and accelerometer was based on the paucity of ferrous or magnetic components, ensuring the MRI compatibility. However, the tablet had some ferrous components and at close proximity to the MRI some pull was noted, so the tablet was kept out of the 200-gauss field.

Spiralgrams were collected before the procedure in the preparation area, then in the MRI before any sonication, and subsequently after each sonication. Finally, a post-procedure spiralgram was also obtained after removal of the head frame. For each timepoint, every patient also completed standard paper spirals in addition to tablet/accelerometer-based assessments. Off-line analysis of cohort data was performed using MATLAB (MathWorks, Natick, MA, USA).

### Data Analysis

The quantitative improvement for the spiral drawing task during the MRgFUS procedure was analyzed in four ways: accelerometer analysis (as measured by the accelerometer), drawing analysis (computerized detection of tremor from image of spiral drawing), combined analysis (the mean of the two previously listed methods), and TETRAS score (neurologist rating of the spiral drawing).

Accelerometer data is transmitted to the application in terms of accelerations in the x, y, and z planes. The motion in the three planes were summed, then filtered using a bandpass filter with a passband of 2-14Hz. Time segments without movements are automatically removed. The power spectral density (PSD) of the filtered signal was obtained using Welch’s PSD algorithm with 2-second segments and 50% overlap between segments. For each recorded spiral, the metrics used to quantify the tremor improvement are a combination of the average amplitude of the filtered timeseries, the area under the curve of the filtered timeseries, the peak value of the PSD, and the area under the largest peak in the PSD. Tremor improvement was quantified as the percent improvement in the combined metrics. Processing pipeline for the accelerometer-derived data is summarized in Figure 1A.

**Figure 1.**
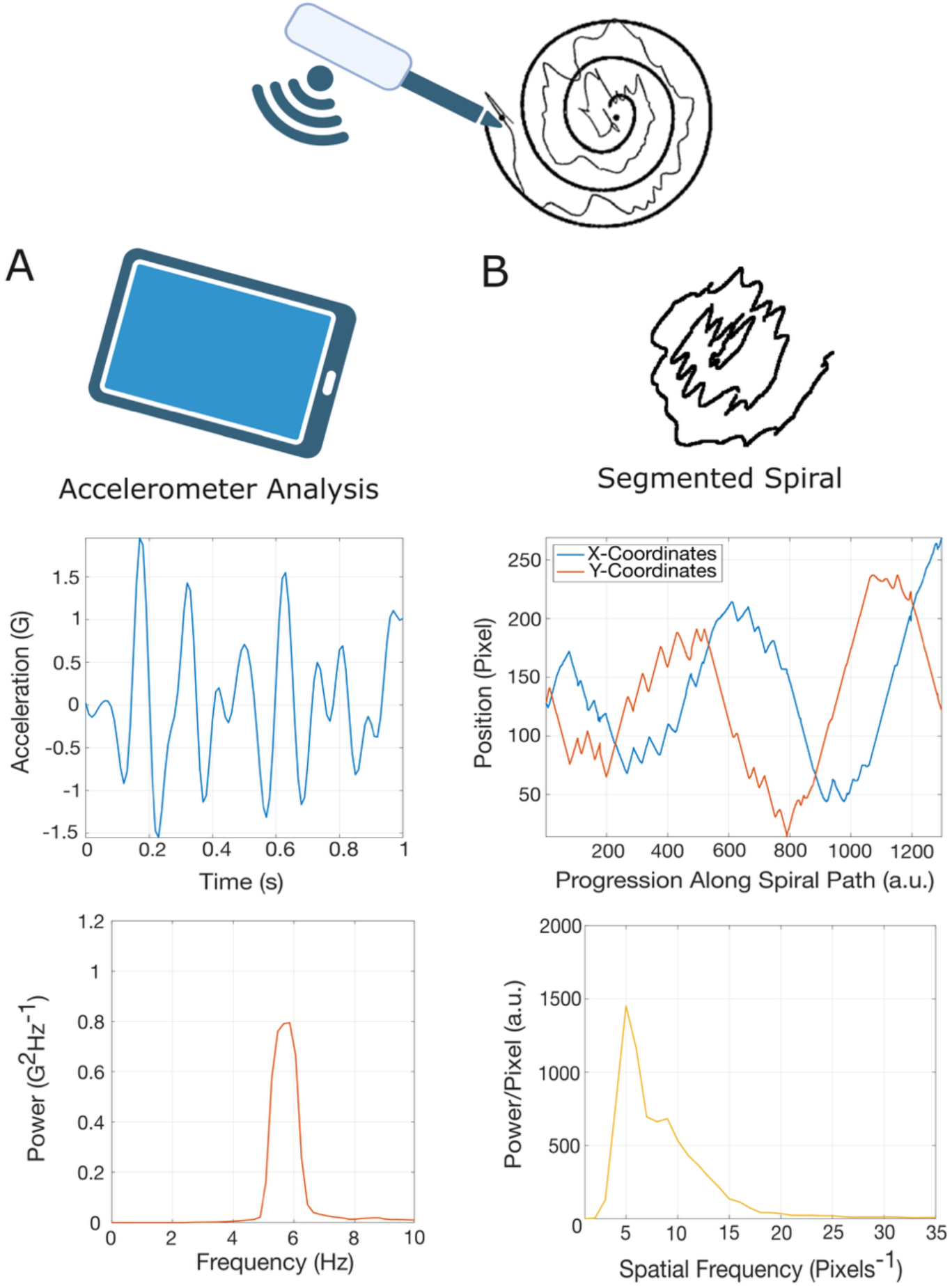
Data acquisition and processing using (A) the accelerometer device, and (B) the paper analysis algorithm. The patient draws the spiral on paper with an accelerometer attached to the pen. The outputs of the accelerometer and the segmented drawn spiral are filtered and the power spectral densities displayed.

The second method of assessing improvement was through the spirals drawn on the tablet or on paper. To analyze the spirals drawn on paper, the template Archimedean spiral was removed from the patient’s drawing (Fig 1B). The spiral was then unraveled to create a linear representation of the spiral drawing, which displayed the relative contributions of the tremor in the x- and y-direction. To unravel the spiral, the central point of the spiral was selected as the start point, and a nearest neighbor search was used to find the path of the spiral. The x and y components of the unraveled spiral were filtered using a high-pass filter with a cutoff frequency of 20/N, where N is number of points in the unraveled spiral. This filter removes components with spatial frequency under 20 cycles per spiral length. To analyze drawings made on the tablet, the process was similar but simplified, since the temporal sequence of the drawing is recorded as part of the task. The spirals on the tablet are similarly unraveled and high-pass filtered. Similar to the accelerometer data, the PSD was computed of the ordered spiral coordinates using Welch’s algorithm with eight evenly divided segments of the input array and 50% overlap. This method provided a measure of the power of the spatial component of the tremor. The metrics used to measure the improvement in tremor intensity from spiralgram drawings were the peak of the spatial PSD and the area under the curve of the spatial PSD. Similar to the accelerometer measurements, percent change in these metrics were computed from baseline, and tremor improvement consisted of a combination of the aforementioned metrics. The third method of analysis was a combination of the accelerometer and the drawn spiral analysis, where the improvement of the two methods was averaged.

Finally, all spirals were evaluated retrospectively by two fellowship-trained movement disorder neurologists using the written section of the essential tremor rating scale (TETRAS). The neurologists were blinded to disease group and accelerometer data. Scores were given from 0-4 (0=no tremor, 4=severe tremor), while allowing half points, as described in guidelines published by Ondo et al (25). The mean score was taken of the two raters for each spiral.

### Statistical Analysis

All statistics were computed using Prism (GraphPad, San Diego, CA, USA). For the pre-vs. post-procedure comparisons, repeated measures one-way analysis of variance (ANOVA) with Tukey’s multiple post-hoc comparisons was performed. For the intra-procedure reported statistics, a mixed-effects analysis using a stacked model, assumed sphericity, and an α = 0.05 was used. A *p-value* of less than 0.05 was considered statistically significant. Statistical tests comparing the tremor rating methods were done in a paired fashion. For the post-procedure improvement comparisons, a mixed-model using a stacked model, assuming sphericity, and an α = 0.05 with Sidak’s multiple post-hoc comparisons was used. Finally, comparisons of the improvement vs. temperature were done using an ordinary one-way ANOVA with Tukey’s multiple post-hoc comparisons. Results are presented and graphed as mean ± SEM unless otherwise denoted.

## Results

### Analysis of tremor characteristics across treatment

The average number of total sonications during the MRgFUS procedure across all patients was 7.4 ± 1.86 (SD) with a range of 4-12 sonications. For each computed metric of improvement with the accelerometer, analyzed spirals (pre-, mid-, and post-procedure) were compared to the pre-sonication baseline and were available instantaneously for digital visualization. The results from a sample patient are shown in Figure 2, showing spiral and accelerometer recordings before, during, and after the procedure.

**Figure 2.**
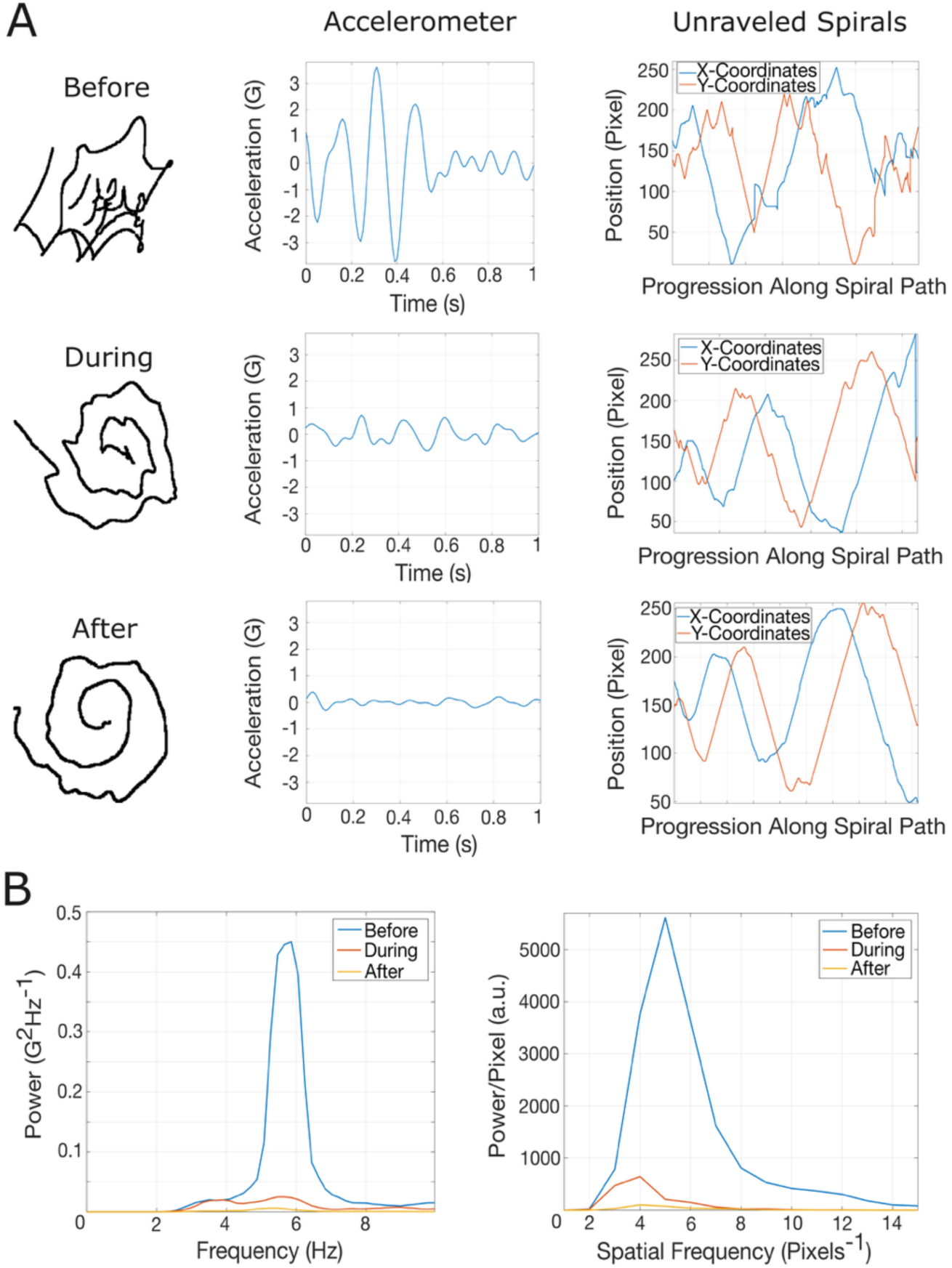
Improvement measurement in a sample patient. A.) Drawn patient spirals, accelerometer readings, and unraveled spirals before, during, and after treatment. B.) Power spectral density in time of the accelerometer and in the spatial domain for the unraveled spiral.

Comparisons were also made across all patients to determine the percent improvement as a result of MRgFUS thalamotomy. The average TETRAS score before the procedure was 3.2 ± 0.12, and after the procedure 2.1 ± 0.13, corresponding to an improvement of 33.6 ± 3.1% (Figure 3A). The average post-procedure improvements for the accelerometer analysis, drawing analysis, combined analysis measured across the different methods were 63.6 ± 7%, 72.4 ± 9.9%, and 68 ± 6.7%, respectively (Figure 3A). One way ANOVA between the four assessment methods showed a significant difference (p<0.001). Post-hoc comparisons showed significant differences between TETRAS ratings and accelerometer, drawing and combined analyses with p values of p=0.0006 and p=0.0003 and p<0.0001, respectively.

**Figure 3.**
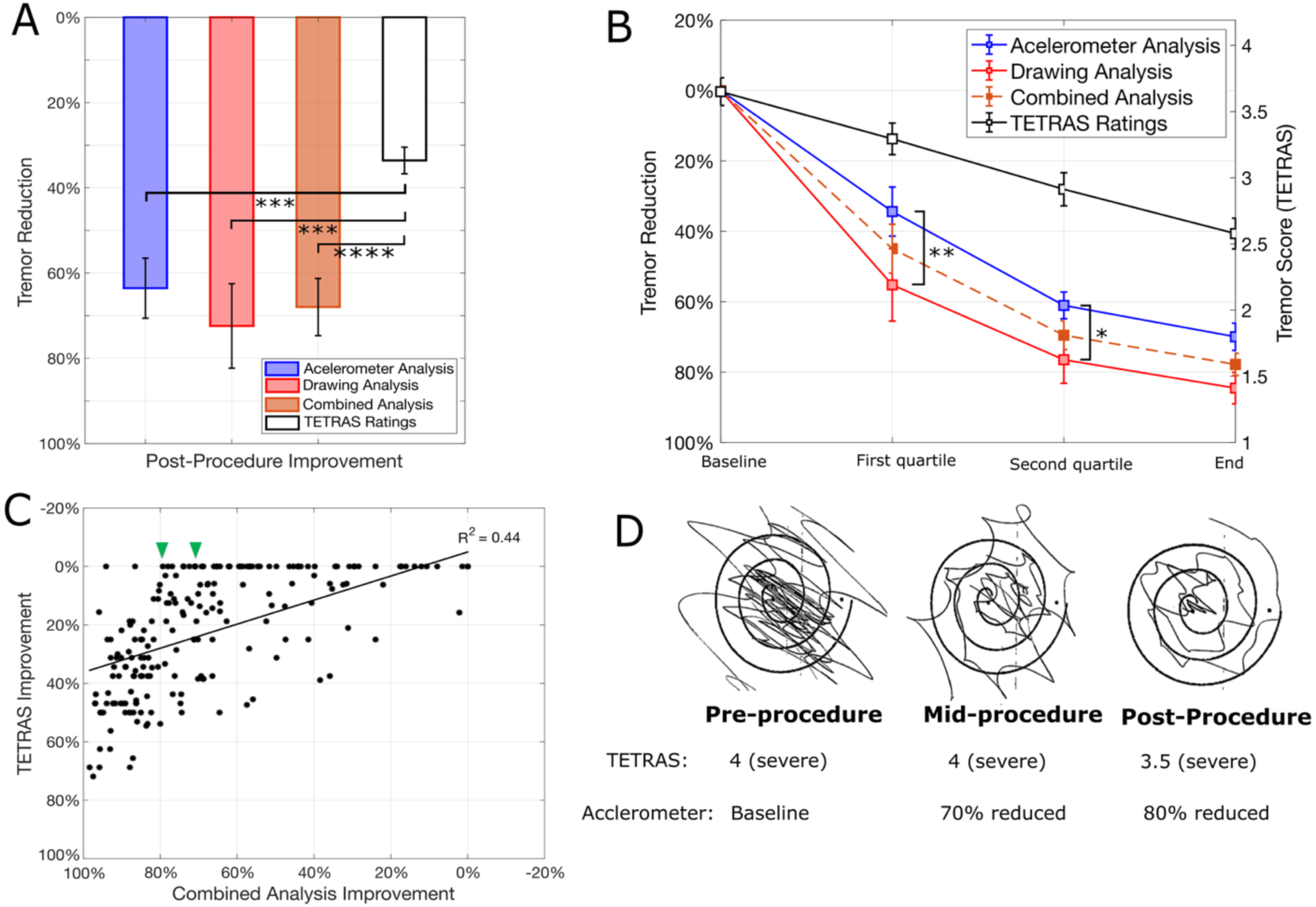
Results across all patients. A.) Post-procedure improvement. B.) Intraprocedural improvement. C.) TETRAS improvement vs. accelerometer improvement. Green arrows denote notable improvement on the accelerometer, however no improvement by TETRAS ratings. D.) Visual example of such spirals.

To assess intraprocedural improvement, the average results of each of the four methods were compared at four different time points, graphically represented in Figure 3B. The label “Second Quarter” represents the mean improvement of sonications 25-50% through the procedure, “Third Quarter” is the mean improvement 50-75% through the procedure, and “End of Procedure” is the mean improvement of the last 75-100% of sonications.

Two-way ANOVA between the accelerometer analysis and drawing analysis methods showed significant differences in both analysis method and time (p<0.001 and p=0.005, respectively). In post-hoc comparison, the drawing analysis and the accelerometer analysis methods showed differences during the procedure in the first and second quartiles (Figure 3B; p=0.001 and p=0.03, respectively), but neither metric was found to be superior (p>0.05) at predicting the final tremor improvement (denoted “End” on Figure 3B).

### Comparison of combined analysis and TETRAS

Percent total improvements from the combined accelerometer and writing analysis and from the TETRAS ratings were plotted in a pairwise fashion (Figure 3C). The calculated Pearson’s correlation indicated moderate positive correlation between the methods (R^2^ = 0.44). On close examination, the curve is left-skewed, and multiple points can be found where TETRAS showed a 0% improvement despite substantial improvement on the combined analysis by as much as 80% (Figure 3D). This finding suggests greater sensitivity of the accelerometer and combined analysis method for assessing improvement of the tremor.

### Percent improvement as a function of sonication temperature

Sonication temperatures ranged between 41°C and 58°C. Improvement is reported within three different temperature ranges: low (< 50°C), medium (50-53°C), and high (>53°C). These temperature ranges were selected because they can guide the expected therapeutic benefit; low temperature for guiding targeting, medium temperature for confirming location and ensuring minimal side effects, and high temperature for the permanent lesioning. The low-temperature lesion showed improvement of 22.8 ± 4.4%, the medium temperature lesion improved by 48.9 ± 5.5%, and the high temperature by 70.8 ± 2.9% (Figure 4). One-way ANOVA showed significant differences between each of the three temperature groups, with p<0.001 for all comparisons.

**Figure 4.**
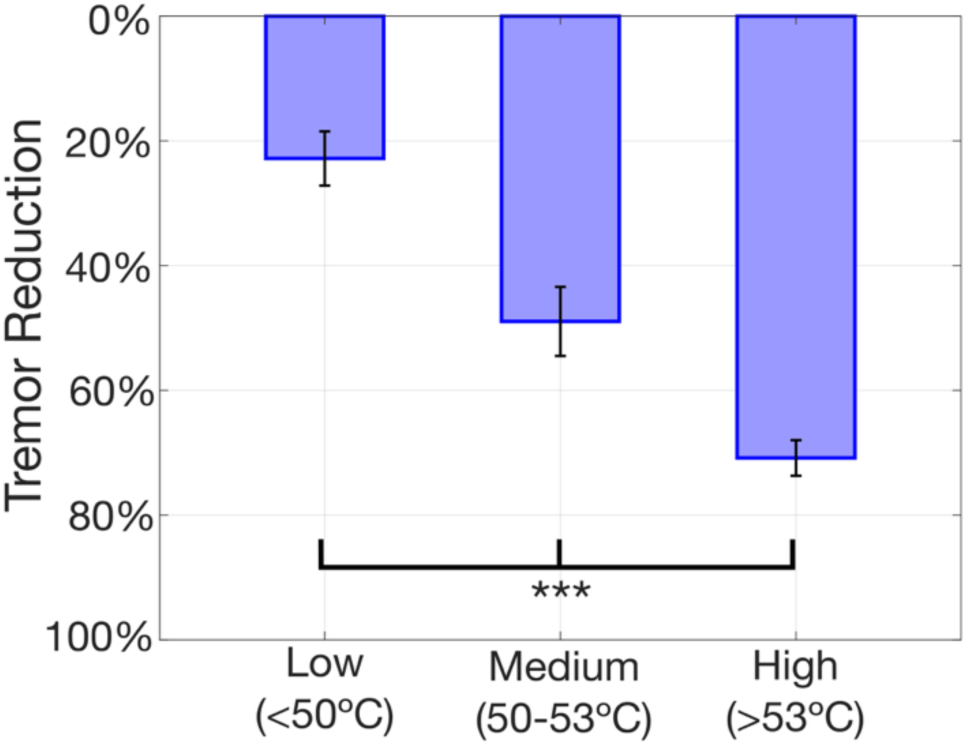
Percent improvement in tremor as a function of sonication temperature, computed as a mean of the accelerometer and drawing analysis methods. Range of sonication temperatures was 41-58°C.

## Discussion

This study demonstrates the potential of using real-time, tablet-based evaluation of spiralgrams through accelerometer and digital writing analysis. The method is low-cost – less than $500 total for a Surface Pro 3 tablet, accelerometer, stylus, and 3D-printed accessories - and easily implemented, and provides a granular degree of measurement of improvement of tremor during focused ultrasound. The schematics and code are open-source and freely available, allowing researchers and community members to easily build and expand on this technology. Because the method uses only an accelerometer and a tablet, it could also be used in deep brain stimulation programming for essential tremor and even assessment of tremor in a movement disorders clinic.

The post-procedural improvements were on average 63% from the accelerometer analysis and 72% from the drawing analysis (68% for the combined accelerometer analysis and drawing analysis). The continuous tracking and analysis of tremor characteristics using the accelerometer and spiral analysis provides a more granular and detailed assessment compared to conventional visual evaluations, and allows calculation of expected improvement at different sonication temperatures. With these values of expected improvements, fine adjustments or additional sonications can be made to optimize treatments if the amount of improvement is below the expected value.

### Use of accelerometry to assess tremor improvement

The use of accelerometers to quantify or characterize tremor have been used extensively, and recently interest has arisen with the use of accelerometers with MRgFUS (26–34). Baek et al positioned three accelerometers on the limb being treated in 18 patients, and found an 80-95% improvement in tremor amplitude, as measured with different tasks (27, 28). Similarly, another group placed just one accelerometer on the index finger, which provided measurements of tremor improvement that were corrected with a similar 50% improvement on the Clinical Rating Scale for Tremor (CRST) using the average power in 1-7Hz band (26). In both cases, the accelerometers were designed as wired devices made by BIOPAC (BIOPAC Systems Inc), which is associated with a higher cost than our system, and also requires the custom BIOPAC data acquisition hardware and software as well as MATLAB software to run the analysis package. Though MATLAB was used to refine figures in this publication for quality, the device itself runs a light-weight and open-source Python based acquisition and analysis software. Additionally, the hardware is self-contained, low-cost, off-the-shelf consumer hardware. Any Windows-compatible tablet and stylus can be used, and the cost of 3D-printing the accelerometer case is negligible. Since all of the code, methods, and model files are open source, the system presented here can be replicated easily and at very low cost, and can be easily updated or improved upon. Additionally, the placement of the accelerometer on the stylus rather than attached to the finger makes recording more efficient, as the patient only has to hold the pen rather than also attaching the accelerometer to the finger.

The combined analysis method (combing accelerometer and drawing analysis) also integrates well into most existing clinical protocols, since the improvement in tremor is evaluated through clinical exam and performance of a spiral or line drawing. A clip also exists on the side of the accelerometer, so that other tasks such as drinking from a cup can be used in conjunction or instead of writing analyses. When the patient is secured to the bed of the MRI after the start of the procedure, the patient must draw the spirals from a supine position. This positioning requires the care team to hold the drawing surface parallel to the patient, above their head. Many patients show worsening of their tremor when drawing supine with an outstretched arm, but whether from the difficulty of the position or the stress of the pending procedure cannot be distinguished. Notably, other patients’ tremors improve in this position, possibly due to relaxing when laying down. Despite the difference in drawing position and ergonomics, the presented results in this manuscript support the ability of this device to gauge subtle changes in tremor improvement over the course of the procedure.

### Comparison to Visual Examination and TETRAS Score

The combined use of the accelerometer analysis and drawing analysis produced consistent, objective measures of tremor improvement. TETRAS scoring is limited by its categorical nature, offering only nine distinct levels of severity. In contrast, the accelerometer-based and drawing analysis methods provide a continuous scale of measurement, allowing for finer distinctions in tremor improvement. Continuous and fine measurements can detect small-scale incremental changes that may be missed with the TETRAS ordinal scoring, as noted during the early stages of the procedure, when improvements of only 10-20% are commonly observed. In those cases, the combined analysis was sensitive enough to detect even subtle changes, whereas TETRAS scores remained unchanged. The limitations of TETRAS were evident, as multiple patients maintained a score of “4” across several initial spirals despite showing some limited (∼20-30%) improvement on computer-based metrics. As the procedure progressed and more substantial tremor relief was achieved after several sonications, the TETRAS scores eventually reflected these changes, despite significant functional relief for the patient.

In the same vein, visual inspection for patients with severe tremors also has limitations. Those patients cannot keep the pen on the paper to fully complete the spiral, therefore limiting the reliability of spiral visual assessment. These constraints are most notable during the critical early phases of the procedure, where noting subtle improvements plays a large role in selecting the target for higher energy sonications. Therefore, a more sensitive method of tremor quantification can significantly improve proper alignment and facilitate clinical decision-making.

### Temperature-Dependent Improvement as a Guide for Targeting

The range of expected improvement versus sonication temperature ranges could provide essential insights for optimizing procedure outcomes. Here, we show that the degree of improvement correlates well with the temperature achieved, with highly predictable range across multiple measurements. Therefore, if a given sonication does not produce the expected improvement, precise quantification using accelerometer device could prompt early re-evaluation of the targeting. Given the lack of a specific radiological landmark for the VIM nucleus and reliance on average measurements, having a framework based on expected improvement at certain temperatures could enhance targeting precision and improve patient outcomes.

### Limitations

One limitation to our study is the lack of a validated, easily-accessible clinical tool for quantifying tremor, which limits the degree to which this method can be validated. Currently, most clinical evaluations rely on subjective measures, such as visual inspection and TETRAS scoring. TETRAS and the CRST are both well-established and have high inter-rater consistency, and are designed to also include assessments of head tremor, standing tremor, and ability to dress oneself (35, 36). While excellent at comprehensive tremor assessment, these metrics are not compatible with MRgFUS workflow. The combined analysis described here provides a much-needed metric focusing on the upper extremity tremor alone, which can be specifically used to assess tremor during MRI-based procedures. Nevertheless, the presented data are preliminary, and additional testing is needed to fully establish the capabilities and limitations of this analysis method. Since tremor severity may vary considerably within a short time period, presumably due to variations in stress, pain, body temperature, or anxiety, the consistency of measurements must be further validated. Finally, further investigation across providers and different institutions is needed to evaluate the external validity of measurements.

## Conclusion

This study supports the use of a low-cost, quantitative tremor analysis method of upper extremity tremor detection. By combining accelerometry and drawing analysis, this method provides objective assessment of upper extremity tremor improvement when used during MRgFUS treatment of ET and tremor-dominant PD. We have demonstrated its utility in providing continuous improvement estimates that can supplement visual evaluations and enhance clinical decision-making. Future work on the device will be aimed at refining and validating the analysis methods to ensure they are fully in accord with clinical assessment. Future studies will focus on validating findings presented here with larger, more diverse patient cohorts across multiple institutions, and determining the effect this tool has on patient-reported outcomes.

## Data Availability

All data produced in the present study are available upon reasonable request to the authors.

https://github.com/clearylabOHSU/tremor_analysis

## Acknowledgements

DC, AR conceptualized the study. DC, AT designed the study. AT, DC, AR collected the data. AT, DC, LN, DS, NU, BS, MH, MJ, HS performed the analysis. AT drafted the initial manuscript. AT, DR, NU, DS, LN, AR, MH, BS, MH, MJ reviewed and edited the manuscript. DC supervised the project and acquired funding. All authors read and approved the final version of the manuscript.

Funding from Oregon Medical Research Foundation (Project #1029114). This work was also supported by the Veterans Affairs (VA) Clinical Science Research and Development (IK2 CX00253) and the Parkinson’s Disease Research, Education, Clinical Center (PADRECC) at the VA Portland Health Care System.

## Supplemental Information

**Supplemental Table.** Intraprocedure Improvement.

|  | Pre-procedure | Second Quarter | Third Quarter | End of Procedure |
| --- | --- | --- | --- | --- |
| **Accelerometer Analysis** | — | 34.4 ± 7% | 61.1 ± 3.8% | 70 ± 3.9% |
| **Drawing Analysis** | — | 55.2 ± 10.2% | 76.5 ± 6.7% | 84.5 ± 4.5% |
| **Combined Analysis** | — | 44.9 ± 6.9% | 69.5 ± 4.1% | 77.8 ± 3.2% |
| **TETRAS Ratings** | — | 0.3 ± 0.5% | 8.8 ± 2.4% | 27.9 ± 2.8% |
| **TETRAS Score (Raw)** | 3.65 ± 0.1 | 3.29 ± 0.12 | 2.91 ± 0.13 | 2.58 ± 0.12 |

Improvement measured using accelerometer analysis, drawing analysis (both percent change), and TETRAS score, as rated by two neurologists (score between 0-4). Information is noted as mean ± standard error of the mean.

## Notes

**Funding**: Funding from Oregon Medical Research Foundation (Project #1029114). This work was also supported by the Veterans Affairs (VA) Clinical Science Research and Development (IK2 CX00253) and the Parkinson’s Disease Research, Education, Clinical Center (PADRECC) at the VA Portland Health Care System.

### Competing Interest Statement

The authors have declared no competing interest.

### Funding Statement

This work received funding from Oregon Medical Research Foundation (Project #1029114). This work was also supported by the Veterans Affairs (VA) Clinical Science Research and Development (IK2 CX00253) and the Parkinson's Disease Research, Education, Clinical Center (PADRECC) at the VA Portland Health Care System.

### Author Declarations

Ethics institutional review board of Oregon Health and Sciences University gave ethical approval for this work (IRB #00026630).

